# Transcriptomic evidence linking adaptive immunity and the IGF-1 pathway in carpal tunnel syndrome

**DOI:** 10.1101/2025.11.17.25340409

**Authors:** Azzurra Laura De Pace, Georgios Baskozos, Annina B Schmid, Dominic Furniss, Akira Wiberg

## Abstract

Carpal tunnel syndrome (CTS), the most common entrapment neuropathy, is characterised by fibrosis and thickening of the subsynovial connective tissue (SSCT) surrounding the median nerve. Although traditionally considered “non-inflammatory,” emerging evidence indicates immune involvement, including elevated cytokines and T-lymphocyte infiltration within the SSCT. Insulin-like growth factor 1 (IGF-1) has been implicated as a potential driver of fibrosis in CTS. Genome-wide association studies identified rs62175241 as a shared risk locus for CTS and trigger finger, where the protective T allele upregulates the long non-coding RNA *DIRC3* and its downstream target *IGFBP5*. Increased IGFBP5 suppresses IGF-1 signalling by binding the ligand, and in other fibrotic diseases has also been linked to T-cell regulation, suggesting a dual fibrotic and immunomodulatory role in CTS.

To investigate the link between fibrosis, inflammation, and the IGF-1 pathway in CTS, we performed bulk RNA-sequencing on SSCT from CTS patients stratified by genotype at the *DIRC3* locus. Differential expression analysis of high-risk versus intermediate- and low-risk genotypes at the *DIRC3* locus revealed 32 upregulated and 316 downregulated genes in high-risk individuals. Upregulated genes included metabolic regulators (*ADIPOQ, GPD1, KLB*), whereas downregulated genes encompassed immune mediators (*CXCL11, MMP9, IL4I1*).

Downregulated genes were enriched for pathways related to adaptive immune responses, including T-cell regulation, challenging the prevailing model of strictly non-inflammatory fibrosis. Furthermore, several components of the IGF axis (*IGFBP5*, *IGFLR1*, *IGF2BP3*) were downregulated in high-risk patients, supporting a role for IGF signalling in CTS. These findings provide evidence to support a model in which dysregulation of IGF-1 signalling intersects with adaptive immune responses to drive fibrosis in CTS, challenging the traditional view of the disease as purely non-inflammatory.

## Introduction

Carpal Tunnel syndrome (CTS), caused by compression of the median nerve in the wrist, is the most common entrapment neuropathy worldwide, affecting around 2-5% of the adult population^1^. Patients experience pain, tingling, numbness, and eventually wasting of the thenar muscles of the hand, leading to functional impairment^2^. CTS is one of the leading causes of work disability and has a significant socioeconomic burden. In the United States over 600,000 carpal tunnel decompression surgeries are performed annually with an estimated economic cost of $5 billion per year^3–5^. However, in up to 25% of cases, symptoms persist even after surgery^6^.

Despite being such a common condition, the aetiology of CTS is relatively under-studied and poorly understood. Most commonly, CTS is believed to arise because of increased pressure in the carpal tunnel which leads to impaired blood supply to the median nerve subsequently causing demyelination and axonal loss^7,8^. The subsynovial connective tissue (SSCT) is a connective tissue layer found between the flexor tendons and the bursa which aids gliding movements between the tendons and the median nerve^8,9^. The prevailing pathophysiological model of CTS involves injury to the SSCT, resulting in fibrosis and hyperplasia of this tissue, which eventually leads to nerve tethering and ischemia^9^. This fibrosis of the SSCT has historically been defined as “non-inflammatory”^10,11^, and is characterised by deposition of excess extracellular matrix (ECM) driven by fibrotic growth factors like TGF-β and PDGFR^10,12–14^.

The long-standing “non-inflammatory” model of CTS pathogenesis, based primarily on basic histological studies^10,13–15^, has been increasingly challenged by evidence suggesting that inflammation is the driver of these pathological changes^16–18^. Notably, we have previously demonstrated elevated levels of CD3+ T cells within the SSCT in patients with CTS^17^. Furthermore, patients with CTS have increased circulating levels of cytokines such as CCL5 and CXCL8^16^, alongside upregulation of CD4+ memory T cells^16,17^.

We previously undertook a genome-wide association study (GWAS) to identify shared genetic risk variants between CTS and trigger finger, a disease which commonly co-occurs with CTS and is also characterised by fibrosis of the connective tissue. This work revealed the existence of a protective T allele (rs62175241) at the *DIRC3* locus on chromosome 2^3^. This variant results in increased expression of long non-coding RNA *DIRC3* and its target IGF-binding protein 5 *(IGFBP5*)^3^. Members of the IGF binding protein family modulate the activity of insulin-like growth factors (IGF) by binding to and regulating the bioavailability of IGF-1^19,20^. Notably, circulating levels of IGF-1 were found to be elevated in patients with CTS^3^. It was therefore hypothesised that in carriers of the protective allele, upregulated IGFBP5 may attenuate IGF-1 signalling by sequestering the IGF-1 ligand^3^. Beyond its canonical role in cell growth and differentiation, the IGF axis has also been implicated in immune homeostasis by maintaining balance between different T-cell subsets^21^. The link between IGFBP-5 and immune regulation has also been shown in other models of disease, including lung fibrosis and colitis^22,23^.

In summary, clinical studies on CTS pathophysiology have implicated multiple biological pathways including ECM hyperplasia and immune dysregulation. To gain further insights into the role of the IGF-1 axis in CTS, we ran secondary analysis on an existing RNA-seq dataset of SSCT samples stratified by *DIRC3* genotype. Our objective was to define the transcriptomic differences associated with high- and low/intermediate-risk alleles and to clarify the interplay between fibrosis, immunity, and IGF-1 signalling in CTS.

## Methods

### Patient cohort

There were 84 participants of whom 31 were male with a mean age of 66.8 (±11.9) years, and 53 female with a mean age of 60.0 (±13.7) years, with CTS. All participants had a clinical diagnosis of CTS, and underwent surgical treatment at the Nuffield Orthopaedic Centre, Oxford University Hospitals NHS Trust. The participants were recruited to two ethically-approved clinical studies, described previously^3,24^: the Pain in Neuropathy Study (PiNS; 10/H07056/35), and the Molecular Genetics of Carpal Tunnel Syndrome (MGCTS) study (16/LO/1920). The CTS cases from the PiNS study had clinically and electrodiagnostically confirmed CTS and were recruited from the surgical waiting lists at Oxford University Hospitals NHS Foundation Trust. The CTS cases from the MGCTS cohort were recruited through hand surgery clinics in Oxford at the time when they were listed to undergo carpal tunnel decompression surgery. Age, sex, BMI, height, ethnicity was recorded for each participant and are reported in Supplementary Table 1.

### RNA extraction

RNA was extracted as described previously^3,24^. Briefly, patients with clinically diagnosed CTS underwent carpal tunnel decompression surgery, during which SSCT specimens were collected. Samples were preserved in RNAlater (Thermo Fisher, MA, USA) before extraction using the High Pure RNA Isolation Kit (Roche, Basel, Switzerland).

### Library preparation and sequencing

RNA samples were sent to the Centre for Human Genetics at the University of Oxford. RNA-Seq was performed in two batches (batch 1: PiNS, July 2017; batch 2, MGCTS, March 2019). The same sequencing protocol was applied to both batches. Library preparation was poly(A)-enriched and directional, and performed using the Illumina TruSeq Stranded mRNA Library Prep Kit and standard universal Illumina multiplexing adaptors. Conversion of the poly(A)-selected RNA to cDNA was performed using the strand-specific dUTP strand-marking protocol^25^, and amplification used unique dual indexing.

Sequencing was performed using the Illumina HiSeq4000 system with 75 bp read length and paired-end reads.

### RNA-seq analysis

FastQ sequencing files were produced that encode quality metrics following the Sanger standard, i.e. Sanger qualities, using the standard Phred score^26^. All sequencing lanes gave a high-yield and consistent GC content, consistent and expected sequence inserts between the paired-end adaptors and high-quality base calling. Individuals were multiplexed in lanes. Reads were mapped to the GRCh38 human genome (Ensembl release 32) and gene counts were assigned using Salmon (v1.12.4)^27^ in *quasi-mapping* mode with selective alignment enabled to improve quantification accuracy. Gene-level transcript counts were summarized based on Ensembl gene annotations.

The resulting count matrix was imported into R (v4.3.1)^28^ using Tximeta^29^ for downstream analysis. Genes with fewer than 10 counts in at least half of the smallest experimental group were filtered out prior to analysis to reduce noise as described in the DESeq2 documentation^30^. Differential expression analysis was performed using the DESeq2 package (v1.42.0)^30^, controlling for potential confounders by specifying a design formula of ∼ batch + biological sex + CTS_risk_grade, where batch indicates the PiNS or MGCTS study. Independent filtering and log₂ fold change shrinkage were applied as implemented in DESeq2^30^. Shrinkage of LFCs was performed using the *apeglm* method to moderate effect sizes for lowly expressed genes^30^. Gene Ontology (GO) enrichment analyses were conducted using clusterProfiler (v4.10.1)^31^ on upregulated and downregulated gene sets separately.

### Statistics

The statistical and graphical analyses were conducted using R (v4.3.1)^28^. The Wald test was used to determine differences in gene expression with an FDR-adjusted p-value < 0.05. The set of genes with an absolute log₂ fold change ≥ 0.32 (1.25-fold change) and adjusted p-value < 0.05 were considered differentially expressed genes (DEG) and were used for the GO analysis. Graphical representations, including volcano plots and heatmaps, were generated using ggplot2 (v3.5.1).

### Genotypic data

Genotypic data for SNP rs62175241 at the *DIRC3* locus were obtained for each of the 81 patients as described previously^3^. Briefly, DNA was extracted from whole blood samples using the PureLink Genomic DNA Kit (Invitrogen, MA, USA) and paired whole-genome genotyping was performed by the Oxford Genomics Centre at the Centre for Human Genetics using the Infinium Global Screening Array-24 v2.0^3^. Pre-phasing and imputation were performed using EAGLE2 and the PBWT algorithm^32,33^.

For the differential gene expression analysis, 18 individuals with genotype CT were grouped together with one individual with genotype TT and labelled as “low/intermediate risk”, whereas 63 individuals with genotype CC were labelled as “high-risk” individuals. The phenotypic data are summarised in Table 1.

**Table 1:** Baseline characteristics in groups with different genotype.

|  | Total | No of Female (%) | No of Male (%) | No of high BMI (%) | Mean Age |
| --- | --- | --- | --- | --- | --- |
| CT and TT | 19 | 8 (42.1) | 11 (57.9) | 13 (68.4) | 65 |
| CC | 63 | 43 (68.2) | 20 (31.7) | 35 (55.5) | 62 |

### Data availability

RNA sequencing data from the Pain in Neuropathy Study (PiNS) cohort has been reported previously and is available at accession GEO108023 (https://www.ncbi.nlm.nih.gov/geo/query/acc.cgi?acc=GSE108023). The relevant raw data (count matrix and genotype calls at rs62175241) in the expanded Oxford-CTS RNA sequencing cohort (including MGCTS) are provided on Github (https://github.com/samkleeman1/cts_tf/).

## Results

Principal component analysis (PCA) of the bulk RNA-sequencing data did not reveal a marked separation between low/intermediate and high-risk individuals for the *DIRC3* locus, indicating that global transcriptomic differences between the groups was relatively subtle (Figure 1A). Despite this, differential gene expression analysis identified a total of 348 significantly altered genes (Figure 1B), comprising 32 upregulated and 316 downregulated genes in high-risk individuals.

**Figure 1:**
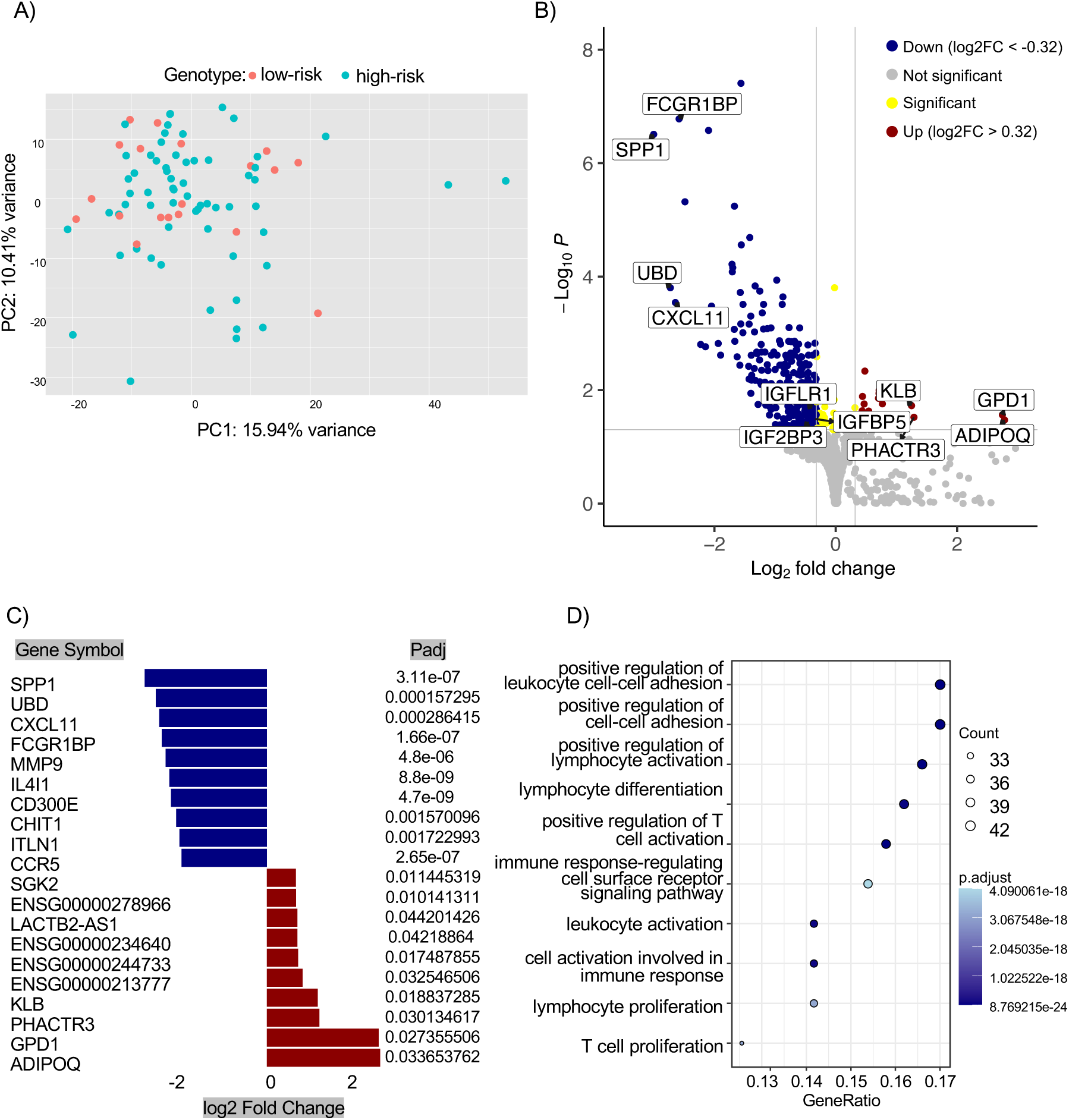
Differential gene expression analysis between different genotypes at the DIRC3 locus (A) Principal component analysis (PCA) of RNA-seq data from subsynovial connective tissue (SSCT) shows no clear global separation between low/intermediate-risk (CT/TT) and high-risk (CC) individuals at the DIRC3 locus. (B) Volcano plot of differential expression analysis (DESeq2) comparing high- vs low/intermediate-risk genotypes. A total of 348 genes were significantly altered (FDR < 0.05), with 32 upregulated (red) and 316 downregulated (blue) in high-risk individuals. (C) Top 10 upregulated and top 10 downregulated genes ranked by log2 fold change. (D) Gene Ontology enrichment analysis of downregulated genes. Abbreviations: DEG, differentially expressed gene; GO, Gene Ontology; SSCT, subsynovial connective tissue.

Among the top upregulated genes (Figure 1C), were genes involved in lipid metabolism and insulin sensitivity (*ADIPOQ, GPD1*)^34–36^, and fibroblast signalling and growth factor modulation (*KLB*)^37,38^. Four long non-coding RNA transcripts (*LACTB2-AS1, ENSG00000234640, ENSG00000244733, ENSG00000278966*) were also significantly upregulated, hinting at broader regulatory shifts in gene networks.

In contrast, the top 10 downregulated genes by fold-change were: *SPP1, CXCL11, UBD, FCGR1BP, MMP9, IL4I1, CHIT1, CD300E, ITLN1 and CCR5* (Figure 1C), all of which are associated with immune regulation and inflammatory signalling. Notably, *SPP1* (*osteopontin*) can act as a cytokine to induce IFN-gamma and IL-12 production^39,40^; CXCL11 acts as a chemokine driving T-cell recruitment to inflamed tissues^41^; and MMP9 is a matrix metalloproteinase with a role in local proteolysis of the extracellular matrix and in leukocyte migration^42,43^. The concurrent downregulation of these immune mediators suggests suppression of local adaptive immune activity in high-risk individuals.

While upregulated genes suggest metabolic changes within SSCT in CTS, pathway enrichment analysis did not identify statistically significant functional categories among the upregulated set. By contrast, Gene Ontology (GO analysis) of downregulated genes in high-risk individuals revealed significant dysregulation of pathways linked to adaptive immune response, including T-cell activation, cytokine-mediated signalling, and lymphocyte differentiation (Figure 1D). These findings indicate that altered *DIRC3* genotype is associated with transcriptional downregulation of immune effector pathways within the SSCT.

Finally, we observed that several genes integral to the IGF-1 signalling pathway were downregulated in high-risk individuals. Specifically, *IGFBP5*, the IGF-like family receptor 1 (*IGFLR1*), and *IGF2BP3* (an RNA-binding protein that stabilizes IGF2 mRNA^44^) were all significantly reduced in high-risk genotype group. Collectively, these transcriptomic changes point towards co-ordinated dysregulation of both immune- and IGF-related pathways, implicating their interaction in the fibrotic pathology of CTS.

## Discussion

This study sought to investigate variation in the transcriptome of CTS patients stratified by genetic risk at the *DIRC3* locus with the aim of clarifying the molecular mechanisms underlying disease susceptibility. Specifically, we aimed to define how genetic variants at this locus influence the interplay between fibrosis, immunity, and IGF-1 signalling. Although principal component analysis did not reveal clear clustering between high and low/intermediate-risk genotype groups, likely reflecting shared disease status among all participants, differential gene expression analysis identified dysregulation of a total of 348 genes. These transcriptomic differences provide novel insights into how variation at the *DIRC3-IGFBP5* axis may modulate both immune and metabolic signalling, thereby contributing to the complex pathophysiology of CTS.

Among 32 genes upregulated in high-risk individuals, *ADIPOQ* or Adiponectin was the most prominently expressed. Adiponectin is secreted by adipocytes and has been associated with various disorders including diabetes, obesity, and cardiovascular disease^35,36^. Given that higher body-mass index (BMI) and metabolic syndrome are risk factors for CTS^45,46^, elevated adiponectin expression could be a mechanistic link between metabolic dysregulation and tissue fibrosis or inflammation in CTS. Four upregulated genes (*LACTB2-AS1, ENSG00000234640, ENSG00000244733, ENSG00000278966*) were RNA genes of unknown function. It is well known that lncRNAs can have regulatory roles; indeed, the *DIRC3* locus itself encodes a lncRNA. LncRNAs are increasingly recognised as modulators of fibroblastic activity; for example, *HOTAIR* regulates synovial fibroblast activation in rheumatoid arthritis^47^.

A key finding of this study was the downregulation of genes central to the adaptive immune response in high-risk individuals, including chemokines (*CXCL11*)^41^, matrix remodelling enzymes (*MMP9*)^42,43^, and immune regulators (*IL4I1, CD300E*)^48,49^. Gene ontology analysis revealed enrichment of pathways associated with T-cell activation and lymphocyte differentiation, consistent with dysregulation of adaptive rather than innate immune mechanisms, suggesting involvement of chronic immune pathways rather than an acute response to injury. These results resonate with prior reports showing increased T-cell infiltration in SSCT and elevated cytokine levels in CTS patient serum^16–18^. The observed downregulation of immune-related genes in high-risk individuals may at first seem counterintuitive, as increased disease risk might typically be associated with inflammation. However, this could reflect compensatory suppression of chronic immune activation or regulatory feedback mechanisms that limit excessive inflammation^50^. Alternatively, reduced expression of immune effectors may represent chronic immune exhaustion or impaired immune surveillance in a chronically fibrotic environment. Future work should assess whether these transcriptomic changes correspond to functional differences in immune cell composition or activity within the SSCT.

Our second key finding concerns the IGF-1 axis, which has been implicated in both fibrosis and immune regulation^19,20,22,23^. Circulating IGF-1 levels are elevated in CTS patients, and IGF binding proteins (particularly IGFBP5) can counterbalance fibroblast-driven IGF-1 activity^3^ by sequestering the ligand, thus attenuating its pro-fibrotic effect. Consistent with this regulatory model, *IGFBP5* expression was downregulated in high-risk individuals. Additionally, two other IGF-related genes, *IGFLR1*, and *IGF2BP3*, were found to be downregulated. The downregulation of *MMP9* is also notable due to its well-established links to IGFBP5 in other disease contexts, including breast cancer and cardiovascular disease^51,52^. There are contrasting reports about whether IGFBP5 suppresses or represses MMP9 activity^51–53^, but our study provides further evidence to suggest that IGFBP5 and MMP9 dysregulation may be observed in patients with CTS, reinforcing the concept that fibrotic and immune mechanisms are interdependent.

Overall, our study demonstrates that genetic variation at the *DIRC3* locus influences transcriptional networks involved in both adaptive immunity and IGF-1 signalling. While the functional consequences of these changes remain to be clarified, they provide a framework for linking genetic susceptibility in CTS to immune modulation and the IGF-1 pathway.

## Limitations

This study has several limitations. First, the absence of non-CTS control tissue limits our ability to distinguish genotype-specific effects from general disease-related transcriptional changes. Obtaining an ideal control tissue is ethically and practically challenging, and substitution with unrelated tissue types (e.g., dermal fibroblasts) would introduce confounding. Second, the use of bulk RNA-seq captures averaged gene expression across multiple cell types, potentially masking cell type-specific contributions; future single-cell RNA-seq could better dissect immune and fibroblast interactions within the SSCT. Additionally, because the protective *DIRC3* allele is rare, grouping of low- and intermediate-risk genotypes into a single category was necessary to achieve sufficient sample size to compare two groups. This approach, while pragmatic, introduces some arbitrariness and may have obscured gradations of genetic effect. Finally, while our study provides correlative evidence, functional experiments will be necessary to confirm causal links between DIRC3 genotype, IGF-1 pathway regulation, and immune function.

## Conclusion

Despite its high prevalence and socioeconomic impact, the molecular pathophysiology of CTS remains poorly defined. Our previous work has linked dysregulation of the IGF-1 pathway to increased genetic risk of CTS, and histological studies have revealed immune activation within fibrotic SSCT. By integrating genetic stratification with transcriptomic analysis, this study provides evidence that unifies these observations. Our findings support a model in which aberrant IGF-1 signalling and altered adaptive immune pathways interact to drive connective tissue remodelling and progressive fibrosis, adding further evidence to challenge the historic view of SSCT fibrosis in CTS as a purely non-inflammatory condition. This work aligns with broader concepts in fibrotic disease biology, where immune dysregulation and growth factor signalling act in concert. These insights may inform the development of future therapeutic approaches targeting IGF-1 modulation and/or immune pathways in CTS.

## Acknowledgements and Funding

The authors would like to thank Dr Michael Ng for his support with the pre-processing of the data, including the alignment of the reads with Salmon. AW is supported by an Arthritis UK Career Development Fellowship (Award Reference 23208), and this work was supported by an MRC fellowship (MR/ N001524/1). DF is supported by the NIHR Biomedical Research Centre, Oxford. (Biomedical Research Centre). AS is supported by a Wellcome Trust Clinical Career Development Fellowship (222101/Z/20/Z) and the NIHR Oxford Biomedical Research Centre.

