## Supplementary Table 1 for "Transcriptomic evidence linking adaptive immunity and the IGF-1 pathway in carpal tunnel syndrome"

Supplementary Table 1: Baseline characteristics of cohort.

| | | | |
|---|---|---|---|
| Number of patients | | | |
| | Total | 84 | |
| | Female | 53 | |
| | | Pre-menopause | 16 |
| | | Post-menopause | 37 |
| | Male | 31 | |
| | White | 81 | |
| | Non white | 3 | |
| | Cohort 1 | 41 | |
| | Cohort 2 | 43 | |
| Average (Mean ± SD) | | | |
| | Age | 62.5 ± 13.4 | |
| | | Female | 60.0 ± 13.7 |
| | | Male | 66.8 ± 11.9 |
| | Height (cm) | 166.6 ± 9.0 | |
| | | Female | 162.2 ± 5.6 |
| | | Male | 174.2 ± 8.7 |
| | BMI | 26.9 ± 5.3 | |
| | | Female | 27.4 ± 5.1 |
| | | Male | 26.0 ± 5.0 |
| BMI breakdown (percentage %) | | | |
| | Underweight | 4 (4.8) | |
| | | Female | 0 (0) |
| | | Male | 4 (100) |
| | Normal | 31 (36.9) | |
| | | Female | 20 (64.5) |
| | | Male | 11 (35.5) |
| | Overweight | 26 (30.9) | |
| | | Female | 18 (69.2) |
| | | Male | 8 (30.8) |
| | Obesity | 23 (27.4) | |
| | | Female | 15 (65.2) |
| | | Male | 8 (34.8) |
